# The Mediating Role of Systemic Inflammation and Moderating Role of Race/Ethnicity in Racialized Disparities in Incident Dementia: A Decomposition Analysis

**DOI:** 10.1101/2023.03.22.23287593

**Authors:** César Higgins Tejera, Erin B. Ware, Margaret T. Hicken, Lindsay C. Kobayashi, Herong Wang, Paris B. Adkins-Jackson, Freida Blostein, Matthew Zawistowski, Bhramar Mukherjee, Kelly M. Bakulski

**Author notes:** **Corresponding Author:** César Higgins Tejera, MD, MPH, MS.

## Abstract

**Background:** Exposure to systemic racism is linked to increased dementia burden. To assess systemic inflammation as a potential pathway linking exposure to racism and dementia disparities, we investigated the mediating role of C-reactive protein (CRP), a systemic inflammation marker, and the moderating role of race/ethnicity on racialized disparities in incident dementia.

**Methods:** In the US Health and Retirement Study (n=5,143), serum CRP was measured at baseline (2006, 2008 waves). Incident dementia was classified by cognitive tests over a six-year follow-up. Self-reported racialized categories were a proxy for exposure to the racialization process. We decomposed racialized disparities in dementia incidence (non-Hispanic Black and/or Hispanic vs. non-Hispanic White) into 1) the mediated effect of CRP, 2) the moderated portion attributable to the interaction between racialized group membership and CRP, and 3) the controlled direct effect (other pathways through which racism operates).

**Results:** The 6-year cumulative incidence of dementia was 15.5%. Among minoritized participants (i.e., non-Hispanic Black and/or Hispanic), high CRP levels (> 75^th^ percentile or 4.57μg/mL) was associated with 1.27 (95%CI: 1.01,1.59) times greater risk of incident dementia than low CRP (≤4.57μg/mL). Decomposition analysis comparing minoritized versus non-Hispanic White participants showed that the mediating effect of CRP accounted for 2% (95% CI: 0%, 6%) of the racial disparity, while the interaction effect between minoritized group status and high CRP accounted for 12% (95% CI: 2%, 22%) of the disparity. Findings were robust to potential violations of causal mediation assumptions.

**Conclusions:** Systemic inflammation mediates racialized disparities in incident dementia.

## Introduction

Dementia is an important contributor to morbidity and mortality in the United States, and a debilitating condition that requires caregiving and support with activities of daily living.^1–3^ The burden of dementia is expected to increase due to demographic changes in the elderly population, lack of early diagnosis, and definite treatment.^4^ Disparities in prevalent and incident dementia have been documented across racialized social groups in the United States.^5,6^ Non-Hispanic Black and Hispanic Americans are more likely to develop dementia than their White counterparts.^5–7^ The increasing burden of dementia will disproportionately affect already vulnerable populations.^8^ In general, minoritized (i.e., historically oppressed and marginalized non-Hispanic Black and/or Hispanic individuals) racial groups are at increased risk of developing chronic conditions,^9–12^ and social models indicate that this susceptibility may be due to a deteriorated physiological function caused by racism (a caste-based system that differentially assigns resources to politically constructed racial group designations).^13–16^

Historically, many studies linking race to health in medicine, public health, and epidemiology have wrongly concluded that disparities in health outcomes are the result of groups differences in phenotypic expressions of race.^17–19^ This ideology is a form of race essentialism that is rooted in racist and colonial systems of oppression. However, historians, sociologists, and social epidemiologists adopted the consensus that racialized categories in the United States are socially constructed through racialization, a process by which individuals are grouped into categories (i.e., racialized social groups) and where access to resources and opportunities are granted or denied.^15,20,21^ Racialization is an important determinant of health disparities^22–24^ as it differentially exposes groups to risk in ways that ultimately influence physiological responses, increasing racialized individuals’ susceptibility to health and disease.^15,25^ Theoretical frameworks such as the weathering hypothesis^13^ and biological embedding^26^ illustrate this pathway by describing how minoritized social groups experience physiological changes as a consequence of persistent marginalization, economic deprivation, and political underrepresentation. Therefore, disparities among racialized groups are the product of racism at interpersonal, institutional, structural, and systemic levels.^25,27^

C-reactive protein (CRP), a marker of systemic inflammation, may capture the impact of racialization on physiological responses. Elevated CRP levels are associated with chronic conditions including cardiovascular disease and adverse cognitive status.^28–36^ Some research has shown that non-Hispanic Black women have the highest levels of CRP in comparison to non-Hispanic White women and men, and even non-Hispanic Black men.^37–39^ Circulating levels of CRP have been associated with higher white matter hyperintensity,^40^ Alzheimer’s disease,^35^ and all-cause dementia,^41^ albeit in studies of homogeneous populations that differ from the racial demographic composition of the United States. In a racially diverse population-based study in the United States, elevated serum levels of CRP were associated with two cognitive domains (memory and verbal fluency) at baseline; but not with the rate of cognitive decline.^29^ In this previous study, the association between CRP and cognition was not modified by race. However, several theoretical social frameworks suggest that systemic inflammation is implicated in the production of both health disparities among racialized groups and in dementia in the overall population.^13–15^ Thus, there is reason to believe that systemic inflammation, via elevated CRP, may be important in linking the downstream effects of racialization (i.e., racialized social categories) to accelerated cognitive aging.^13,26^

This study examines the mediating role of systemic inflammation, and the moderating role of race/ethnicity on disparities in incident dementia in a large, diverse, population-based study.^42,43^ We conceptualize health disparities as the result of racialization,^16,44^ therefore, we use causal mediation-interaction analysis to determine whether self-reported racialized social categories (e.g., as a proxy for exposure to racialization) is an effect modifier of the association between systemic inflammation and incident dementia, as well as to understand if systemic inflammation is a mediating pathway of disparities among racialized groups (i.e., non-Hispanic Black and/or Hispanic vs non-Hispanic White). Therefore, when comparing racialized groups, we are comparing forced membership in a minoritized social group (i.e., racialized non-Hispanic Black and/or Hispanic) as opposed to a more privileged one (i.e., racialized as non-Hispanic White), which captures the impact of racialization on health disparities, and not fictionalized genetic or ancestry differences.^44,45,46^ Moreover, because apolipoprotein E (*APOE*) is associated with lower circulating levels of CRP,^47,48^ and *APOE-ε4* allele carrier status is an important genetic marker for dementia risk, more frequently found in non-Hispanic Black Americans,^49^ we use randomized analogue models to test the robustness of our main mediation analysis.

## Methods

### Study Design

The US Health and Retirement Study (HRS) is a longitudinal study of older adults in the United States.^50^ To ensure representativeness of the national demographic composition, the HRS oversamples non-Hispanic Black and Hispanic participants using a multi-stage probability design.^50^ The initial cohort was formed in 1992 and interviews are conducted every two years. Written informed consent was obtained for all participants at data collection. The University of Michigan Institutional Review Board (HUM00128220) approved this secondary data analysis. Survey data are publicly available (https://hrs.isr.umich.edu/data-products), and genetic data are available through dbGaP (https://dbgap.ncbi.nlm.nih.gov; phs000428.v2.p2) and the National Institute on Aging Genetics of Alzheimer’s Disease Data Storage Site (https://dss.niagads.org/; NG00119).

In 2006, a random half of participants was selected for dried blood spot and biomarker assessment and another half in 2008. For this analysis, we selected participants who provided dried blood spot samples and were cognitively normal at their respective baseline (2006 or 2008). Every two years after baseline measurements, participants underwent cognitive assessments; they were followed up either until their cognitive test results indicated dementia or until the end of a six-year follow-up period (2012 or 2014). We explored the association between circulating CRP and 6-year incident dementia in the overall analytical sample and across three racialized groups: non-Hispanic Black, non-Hispanic White, and Hispanic participants. We used mediation-interaction analysis decomposition to explore the moderating effect of racialized categories on the association between systemic inflammation and incident dementia. We also tested whether systemic inflammation was a mediator of racial disparity in incident dementia between non-Hispanic Black and/or Hispanic participants relative to their non-Hispanic White counterparts. This study adhered to both the Strengthening the Reporting of Observational Studies in Epidemiology (STROBE) guidelines^51^ and the Guideline for Reporting Mediation Analyses (AGReMA).^52^

### Measures

#### Outcome

Cognitive status was evaluated at baseline and every two years through telephone interview. Cognitive test results were recorded in a continuous scale including: 10 word immediate and delayed recall tests, a serial 7 subtraction test, counting backwards, object naming, and recalling the date, president, and vice-president to assess orientation. The scale ranges from 0-27 points, with larger values indicating better cognitive performance. We used the Langa-Weir cut points to classify participants who scored 0-6 as having dementia, 7-11 as having cognitive impairment non-dementia (CIND), and 12-27 as cognitively normal.^53^ For the purpose of our analysis we focused solely on participants who developed dementia over the 6-year study period, from any non-dementia baseline classification. Proxy respondents were not included in our sample, as they did not provide blood spots.

#### Exposure

We used participants’ self-reported racialized categories as a proxy measure of exposure to racialization and compared each minoritized group (non-Hispanic Black and Hispanic) to the most privileged category (non-Hispanic White, reference group).^44^ Because non-Hispanic Black and Hispanic adults undergo cumulative experiences of marginalization and political underrepresentation in the United States relative to their non-Hispanic White counterparts, we combined these two groups into a unique minoritized category to leverage a larger sample size, and compared them to the most privileged social group. Throughput the manuscript, when comparing jointly non-Hispanic Black and Hispanic participants to the most privilege group, we use the terminology minoritized group; otherwise, we specify which racialized groups are being compared.

#### Mediator

Circulating CRP was measured in blood spots using enzyme-linked immunosorbent assay (ELISA).^54^ The CRP assay lower limit of detection is 0.035mg/L, within-assay imprecision is 8.1% and between-assay imprecision is 11.0%. CRP concentrations from dried blood spot samples were linearly related to concentrations in plasma samples (n =87 paired samples, Pearson R = 0.99).^54^ For statistical analysis, we considered CRP in its continuous form. As there are no clinical thresholds for CRP, we dichotomized at the 75^th^ percentile (≥4.57μg/mL) of our analytical sample distribution.

#### Covariates

We included potential confounders of the association between our exposure of interest, racialized groups, our mediator CRP, and incident dementia, with all confounders measured at baseline (**Figure 1**). Sociodemographic confounders were self-reported and included age (continuous, in years, calculated from birth date and interview date), gender (female or male), and education (more than college, college or some college, high school or less). Behavioral confounders included smoking status (current/former or never), alcohol consumption (reported as number of drinks a day when drinks, continuous), and self-reported body mass index (calculated as weight kilograms divided by height in meters squared, continuous). Number of chronic health conditions included high blood pressure (yes or no), diabetes (yes or no), cancer (yes or no), lung disease (yes or no), heart disease (yes or no), stroke (yes or no), psychiatric problems (yes or no), and arthritis (yes or no), and was operationalized in our models as a dichotomous variable (at least 1 condition or no). Genetic information on *APOE*-*ε4* allele carrier status (at least 1 copy or no copy) was obtained from phased genetic data imputed to the worldwide 1000 Genomes Project reference panel. Genotyping and imputation information on the Health and Retirement Study is available elsewhere.^55^ We also included baseline survey wave (2006 or 2008) as a covariate to account for unmeasured differences across waves.

**Figure 1:**
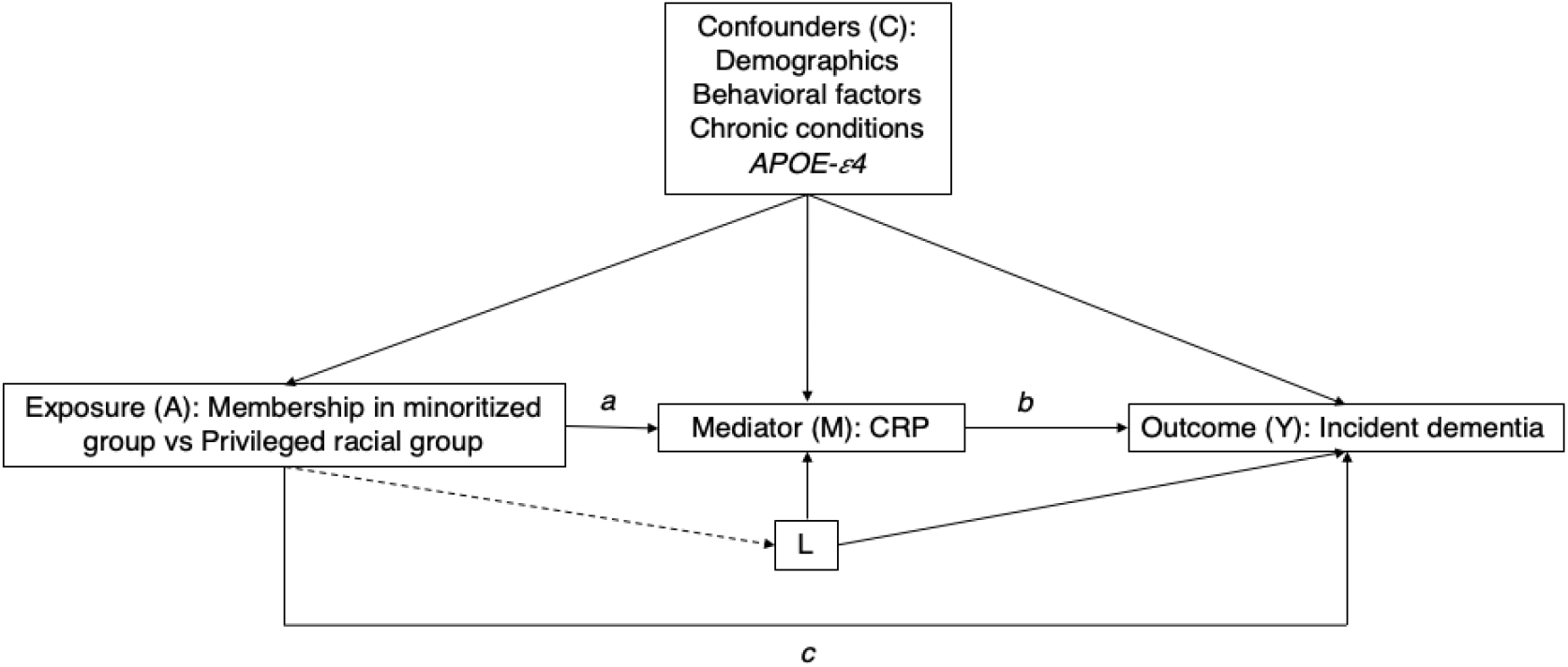
Directed Acyclic Graph illustrating the relationship between racialized social groups, systemic inflammation, and 6-year incident dementia in the Health and Retirement Study. Exposure (A), represents membership in a minoritized or racialized group vs a privileged group (i.e., non-Hispanic Black and/or Hispanic participants vs non-Hispanic White participants). Racialized group membership is directly associated with incident dementia, as denoted in arrow c, and through systemic inflammation (Mediator M) as denoted by arrows (a) and (b). The association between systemic inflammation and incident dementia is denoted by arrow (b). However, the association between systemic inflammation and incident dementia can be modified by membership in minoritized racial status, as this model allows for exposure-mediator interaction. The set of confounders (C) account for exposure (A) - outcome (Y), exposure (A) - mediator (M), and mediator (M) - outcome (Y) confounders. This model also assumes that there is not mediator-outcome confounder (L) affected by the exposure (A)

## Statistical Analysis

Our analytical sample included participants with complete information for all our covariates of interest and those who developed dementia over the 6-year period from any non-dementia baseline classification. We examined the distributions of all baseline covariates by each of self-reported racialized categories, quartiles of the CRP distribution, and incident dementia using bivariate statistical tests, as appropriate. We used kernel density plots to explore the distribution of CRP by the three racialized groups, as well as stratified by *APOE*-*ε4* allele carrier status and racialized categories. Additionally, we explored the distribution of CRP concentrations by racialized groups and self-reported gender categories using frequency statistics. We dichotomized CRP at the 75^th^ percentile of the study sample distribution and categorized those with levels above or equal to the 75^th^ percentile (≥4.57μg/mL) as high, and those less than 75^th^ percentile (<4.57μg/mL) as low. Because our primary endpoint of interest was incident dementia, we excluded participants with CIND who never transitioned to dementia over the 6-year period, and those with incomplete information on covariates of interest (**Supplemental Figure 1**).

In the overall sample and stratified by either minoritized status or racialized groups, we employed multivariable-adjusted Poisson regression models with a logarithmic link function and time-to-dementia as an offset variable, to estimate incident rate ratios of dementia between participants with high CRP (≥75^th^ percentile) versus low CRP. In order to understand how the magnitude of the association between our mediator (CRP) and our outcome (incident dementia) changed with different sets of confounders, we fitted four sequential regression models: an unadjusted model, a demographic model (adjusted for age, gender, education, *APOE*-*ε4* allele status, and survey wave), a behavioral model (demographic adjusted model, and smoking status, alcohol consumption, and body mass index), and a chronic condition model (behavioral adjusted model, and chronic conditions). We employed logistic regression analysis to estimate the association between each of racialized group (each of non-Hispanic Black and Hispanic, versus non-Hispanic White) and minoritized group (non-Hispanic Black or Hispanic, versus non-Hispanic White) and the odds of high CRP levels (≥ 75^th^ percentile), adjusted for the same confounders as described above. We performed four-way mediation-interaction decomposition analysis to evaluate whether CRP mediated disparities among racialized groups in incident dementia using the CMAverse R studio package, accounting for any interaction effect between minoritized group status and CRP.^56^ This interaction effect allowed us to capture whether belonging to a minoritized group differentially affected the strength of the association between systemic inflammation and incident dementia. Analyses were conducted using R statistical software (version 3.6.2) and Stata (v17). A second analyst performed complete code review. Code to produce these analyses is available (https://github.com/bakulskilab)

### Sensitivity Analysis

Mediation analysis assumes that if the adjustment set of covariates is sufficient to control for all exposure-outcome, mediator-outcome, and exposure-mediator confounders, then natural or pure indirect effects are identifiable.^57^ However, an additional assumption to identify indirect effects is needed: no mediator-outcome confounder should be affected by the exposure.^57^ Our directed acyclic graph **(Figure 1)** illustrates a potential scenario in which a variable (L) acts as a mediator-outcome confounder affected by the exposure. In the HRS, self-reported racialized categories are paired to geographic genetic ancestry groups by study design.^55^ Because non-Hispanic Black participants of African ancestry are more likely to be carriers of the *APOE-ε4* allele **(variable L, Figure 1)**, and this allele is associated with both circulating levels of CRP (mediator) and dementia (outcome), we employed randomized analogue models to test the robustness of our mediation analysis findings.^57^ Additionally, because our primary interest is the identification of the mediated effect of CRP on disparities, we calculated mediational E-values to examine the extent to which an unmeasured confounder could explained away our observed mediational effect.^58^ Mediational E-values were calculated for the total natural indirect effect (proportion mediated) for both regression-based and randomized analogue estimates.

## Results

### Sample characteristics

Our analytic sample size included 5,143 participants (**Supplemental Figure 1**). On average, participants were 66.4 years of age, 62% were female, 9.5% were non-Hispanic Black, 7.3% were Hispanic, 67% completed high school education or less, and had average CRP concentrations of 4.18μg/mL (**Table 1**). Excluded participants had higher circulating CRP, were older, more likely to be male, non-Hispanic Black or Hispanic, and had completed high school education or less (**Supplemental Table 1**). Cumulative dementia incidence over the 6-year follow-up was 35% for non-Hispanic Black participants, 27% for Hispanic participants, and 12% for non-Hispanic White participants (**Supplemental Table 2**). The highest mean CRP levels were observed in non-Hispanic Black participants (6.2μg/mL) followed by Hispanic participants (4.3μg/mL) and non-Hispanic White participants (3.9μg/mL) (**Supplemental Table 2 & Figure 2**). Non-Hispanic Black female participants had the highest average concentrations of CRP levels (6.3μg/mL), whereas non-Hispanic White male participants had the lowest (3.4μg/mL) (**Supplemental Table 3 & Supplemental Figure 2**).

**Table 1:**
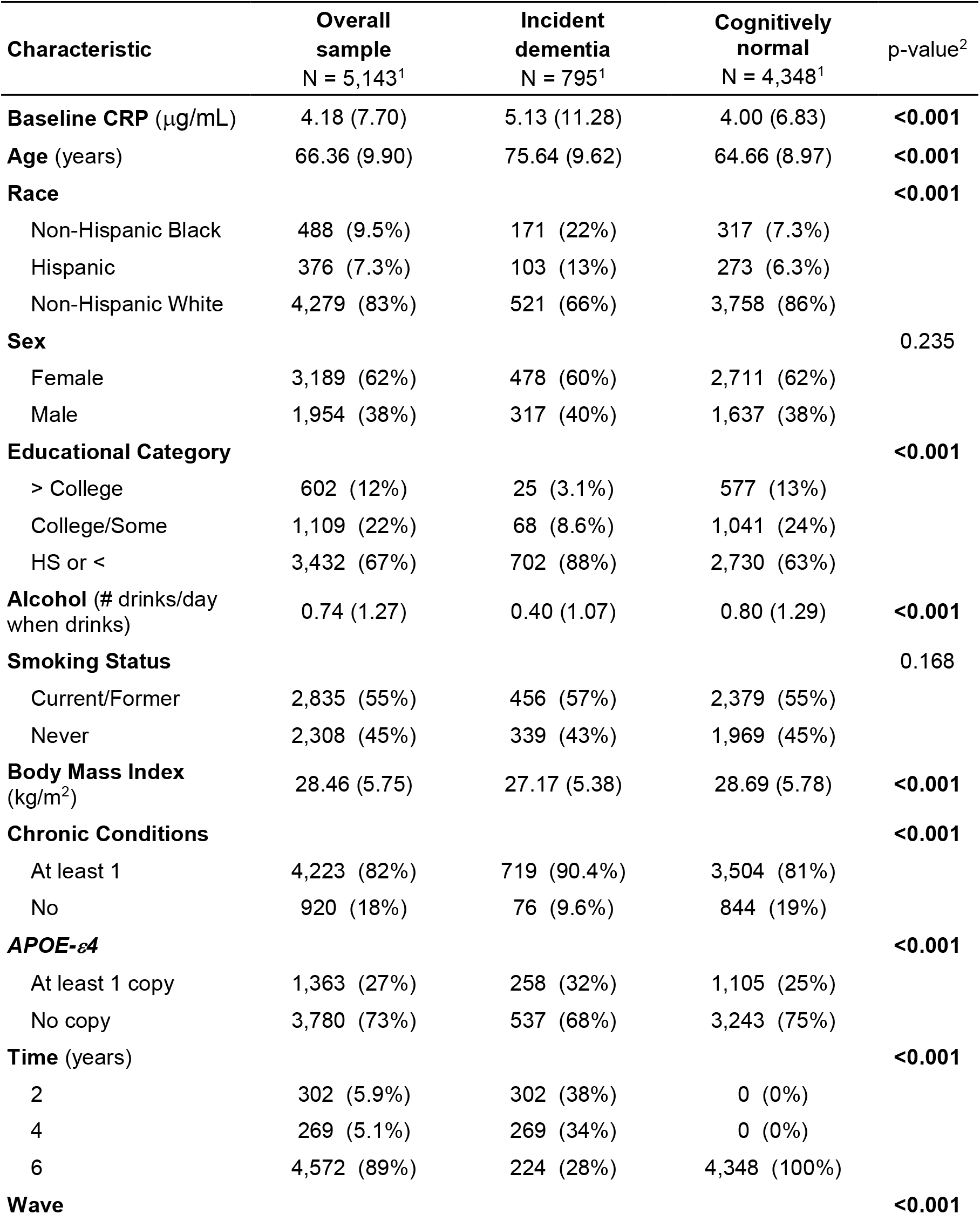
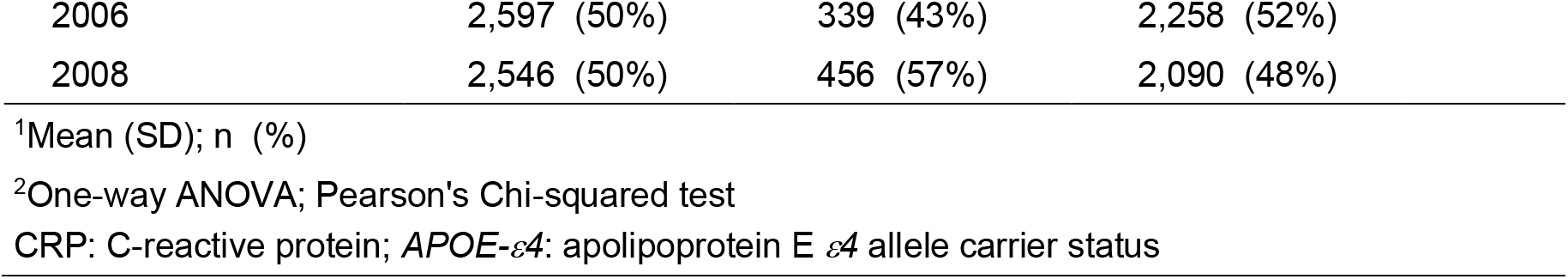
Distribution of baseline sample characteristics by dementia status after 6-year of follow-up, United States Health and Retirement Study, 2006 and 2008

| Characteristic | Overall sample<br>N = 5,143<sup>1</sup> | Incident dementia<br>N = 795<sup>1</sup> | Cognitively normal<br>N = 4,348<sup>1</sup> | p-value<sup>2</sup> |
| --- | --- | --- | --- | --- |
| **Baseline CRP** (µg/mL) | 4.18 (7.70) | 5.13 (11.28) | 4.00 (6.83) | **<0.001** |
| **Age** (years) | 66.36 (9.90) | 75.64 (9.62) | 64.66 (8.97) | **<0.001** |
| **Race** |  |  |  | **<0.001** |
| Non-Hispanic Black | 488 (9.5%) | 171 (22%) | 317 (7.3%) |  |
| Hispanic | 376 (7.3%) | 103 (13%) | 273 (6.3%) |  |
| Non-Hispanic White | 4,279 (83%) | 521 (66%) | 3,758 (86%) |  |
| **Sex** |  |  |  | 0.235 |
| Female | 3,189 (62%) | 478 (60%) | 2,711 (62%) |  |
| Male | 1,954 (38%) | 317 (40%) | 1,637 (38%) |  |
| **Educational Category** |  |  |  | **<0.001** |
| > College | 602 (12%) | 25 (3.1%) | 577 (13%) |  |
| College/Some | 1,109 (22%) | 68 (8.6%) | 1,041 (24%) |  |
| HS or < | 3,432 (67%) | 702 (88%) | 2,730 (63%) |  |
| **Alcohol** (# drinks/day when drinks) | 0.74 (1.27) | 0.40 (1.07) | 0.80 (1.29) | **<0.001** |
| **Smoking Status** |  |  |  | 0.168 |
| Current/Former | 2,835 (55%) | 456 (57%) | 2,379 (55%) |  |
| Never | 2,308 (45%) | 339 (43%) | 1,969 (45%) |  |
| **Body Mass Index** (kg/m<sup>2</sup>) | 28.46 (5.75) | 27.17 (5.38) | 28.69 (5.78) | **<0.001** |
| **Chronic Conditions** |  |  |  | **<0.001** |
| At least 1 | 4,223 (82%) | 719 (90.4%) | 3,504 (81%) |  |
| No | 920 (18%) | 76 (9.6%) | 844 (19%) |  |
| **APOE-ε4** |  |  |  | **<0.001** |
| At least 1 copy | 1,363 (27%) | 258 (32%) | 1,105 (25%) |  |
| No copy | 3,780 (73%) | 537 (68%) | 3,243 (75%) |  |
| **Time** (years) |  |  |  | **<0.001** |
| 2 | 302 (5.9%) | 302 (38%) | 0 (0%) |  |
| 4 | 269 (5.1%) | 269 (34%) | 0 (0%) |  |
| 6 | 4,572 (89%) | 224 (28%) | 4,348 (100%) |  |
| **Wave** |  |  |  | **<0.001** |

| **Characteristic** | **Overall**<br>**sample**<br>**N = 5,143<sup>1</sup>** | **Incident**<br>**dementia**<br>**N = 795<sup>1</sup>** | **Cognitively**<br>**normal**<br>**N = 4,348<sup>1</sup>** | **p-value<sup>2</sup>** |
| --- | --- | --- | --- | --- |
| 2006 | 2,597 (50%) | 339 (43%) | 2,258 (52%) |  |
| 2008 | 2,546 (50%) | 456 (57%) | 2,090 (48%) |  |

<sup>1</sup>Mean (SD); n (%)
<sup>2</sup>One-way ANOVA; Pearson's Chi-squared test
CRP: C-reactive protein; *APOE-ε4*: apolipoprotein E *ε4* allele carrier status

**Figure 2:**
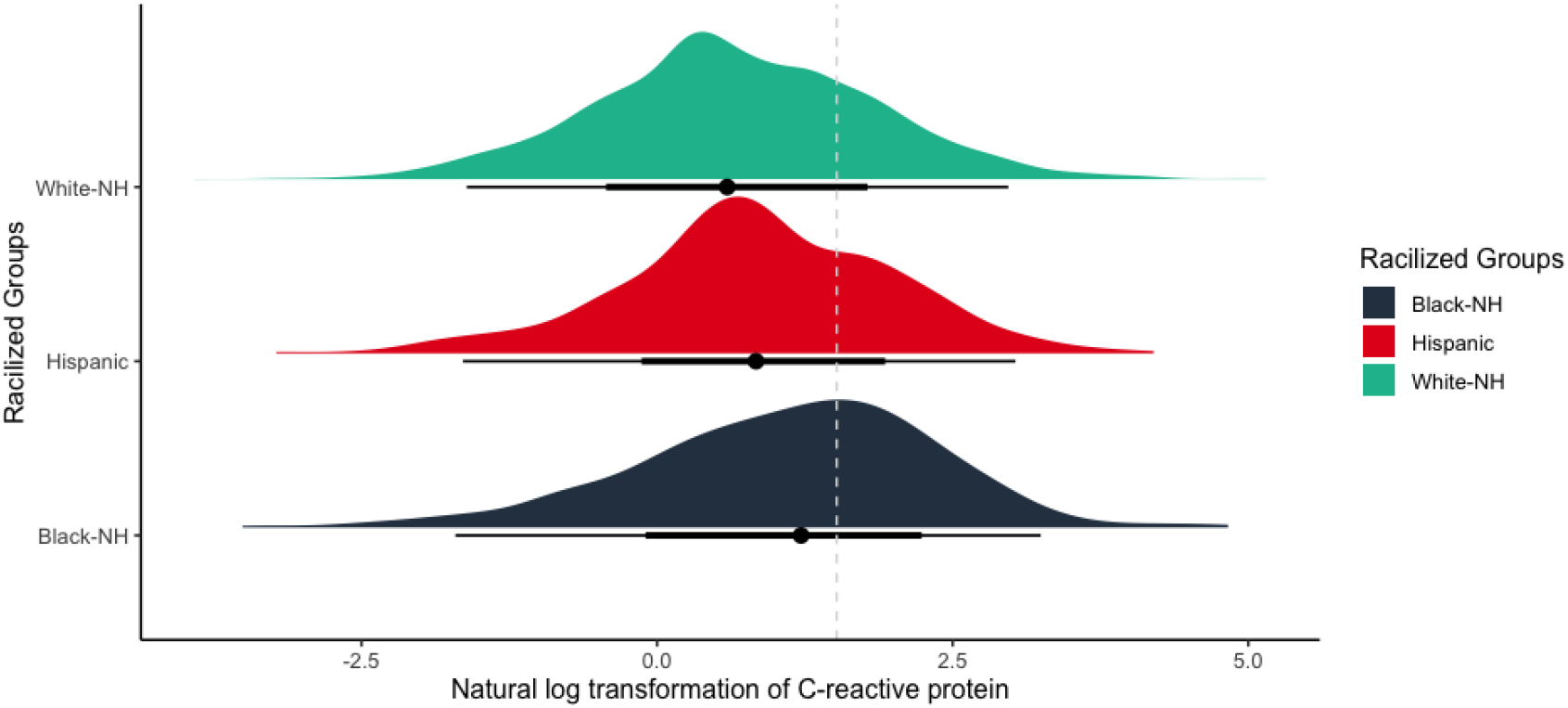
Density plot visualizing the distribution of the natural logarithmic transformation of C-reactive protein (CRP) by racialized social groups in our analytic sample of the Health and Retirement Study. Dotted line denotes the cut off point for elevated levels of CRP at the 75^th^ percentile (>4.57μg/mL). White-NH: non-Hispanic White, Black-NH: non-Hispanic Black.

### Associations between C-reactive protein and incident dementia in the overall analytic sample, by minoritized and racialized groups

On average, participants with incident dementia had higher levels of CRP (5.1μg/mL), compared to participants with normal cognition (4.0μg/mL) (**Table 1**). However, the proportion of incident dementia cases across quartiles of the CRP distribution did not substantially differ. For example, among those with CRP levels >4.57μg/mL (>75^th^ percentile), the 6-year cumulative dementia incidence was 17%, and among those with CRP levels <0.96μg/mL (<25^th^ percentile) was 15% (**Supplemental Table 4**). Participants in the 75^th^ percentile of the CRP distribution were on average of slightly younger age, female, non-Hispanic White, and completed a high school education or less compared to those below the 25^th^ percentile. Additionally, these participants were more likely to be current or former smokers, had fewer drinks per day when they drink, had a larger body mass index on average, were more likely to have at least 1 chronic health condition, and were less likely to be carriers of the *APOE-ε4* allele than those in the lowest quartile of the distribution (<25^th^ percentile) (**Supplemental Table 4**).

In our overall sample, the fully adjusted model showed that among those exposed to high inflammation levels, the 6-year risk of incident dementia was 1.27 (95% CI: 1.10, 1.47) times higher than in those with low inflammation levels (**Table 2**). Sequential adjustment suggested that the strength of the association between high CRP levels and dementia risk increased after conditioning on potential confounders. The association between high CRP levels and incident dementia differed across minoritized and racialized group. For example, among minoritized participants, high CRP was associated with 1.27 (95% CI: 1.01, 1.59) times higher risk of incident dementia than low CRP. Similarly, the risk of 6-year incident dementia for non-Hispanic White participants with high CRP was 1.25 (95% CI: 1.04, 1.50) times higher than those with low CRP. However, among Hispanic participants, high CRP was associated with 1.55 (95% CI: 1.06, 2.27) times higher risk of dementia than low CRP. However, among non-Hispanic Black participants, this association was attenuated, and those with high CRP had only 1.09 (95% CI: 0.83, 1.43) times higher risk of dementia than those with low CRP (**Table 2**).

**Table 2:** Incidence rate ratios from Poisson regression analysis, estimates represent the association between elevated levels of C-reactive protein (CRP) (>4.57μg/mL) and 6-year incident dementia in the United States Health and Retirement Study. Models are stratified by racialized social groups and minoritized status.

|  | Incident Dementia |  |  |  |  |  |  |  |  |  |
| --- | --- | --- | --- | --- | --- | --- | --- | --- | --- | --- |
|  | Overall |  | Minoritized<br>(non-Hispanic Black & Hispanic) |  | Non-Hispanic Black |  | Hispanic |  | Non-Hispanic White |  |
|  | N = 5,143 |  | N = 864 |  | N = 488 |  | N = 376 |  | N = 4,279 |  |
| **Models** |  |  |  |  |  |  |  |  |  |  |
| **Unadjusted** | **IRR<sup>1</sup>** | **95% CI<sup>1</sup>** | **IRR<sup>1</sup>** | **95% CI<sup>1</sup>** | **IRR<sup>1</sup>** | **95% CI<sup>1</sup>** | **IRR<sup>1</sup>** | **95% CI<sup>1</sup>** | **IRR<sup>1</sup>** | **95% CI<sup>1</sup>** |
| < 75<sup>th</sup> | 1 | - | 1 | - | 1 | - | 1 | - | 1 | - |
| $\geq 75^{\text{th}}$ ($\geq 4.57 \mu\text{g/mL}$) | 1.19* | [1.02,1.39] | 1.08 | [0.85,1.36] | 0.84 | [0.62,1.13] | 1.53* | [1.05,2.23] | 1.06 | [0.87,1.29] |
| **Demographic<sup>+</sup>** |  |  |  |  |  |  |  |  |  |  |
| < 75<sup>th</sup> | 1 | - | 1 | - | 1 | - | 1 | - | 1 | - |
| $\geq 75^{\text{th}}$ ($\geq 4.57 \mu\text{g/mL}$) | 1.22** | [1.05,1.41] | 1.26* | [1.01,1.57] | 1.08 | [0.82,1.43] | 1.55* | [1.07,2.26] | 1.17 | [0.97,1.41] |
| **Risk Factors<sup>§</sup>** |  |  |  |  |  |  |  |  |  |  |
| < 75<sup>th</sup> | 1 | - | 1 | - | 1 | - | 1 | - | 1 | - |
| $\geq 75^{\text{th}}$ ($\geq 4.57 \mu\text{g/mL}$) | 1.27** | [1.10,1.47] | 1.27* | [1.01,1.59] | 1.09 | [0.83,1.44] | 1.53* | [1.04,2.24] | 1.25* | [1.04,1.51] |
| **Chronic Conditions<sup>§</sup>** |  |  |  |  |  |  |  |  |  |  |
| < 75<sup>th</sup> | 1 | - | 1 | - | 1 | - | 1 | - | 1 | - |
| $\geq 75^{\text{th}}$ ($\geq 4.57 \mu\text{g/mL}$) | 1.27** | [1.10,1.47] | 1.27* | [1.01,1.59] | 1.09 | [0.83,1.43] | 1.55* | [1.06,2.27] | 1.25* | [1.04,1.50] |

<sup>+</sup> Demographic model: adjusted for age, sex, education categories, *APOE-ε4* allele status, and wave. Note: the demographic model in the overall sample (N=5,143) additionally adjust for racialized social groups
<sup>§</sup> Risk factors model: adjusted for age, sex, education categories, *APOE-ε4* allele status, wave, smoking status, alcohol consumption, body mass index
<sup>§</sup> Chronic conditions model: adjusted for age, sex, education categories, *APOE-ε4* allele status, wave, smoking status, alcohol consumption, body mass index, and chronic conditions
<sup>1</sup> IRR: incidence rate ratio, CI: confidence interval in brackets,
\* $p < 0.05$ , \*\* $p < 0.01$ , \*\*\* $p < 0.001$

### Minoritized and racialized disparities in high circulating levels of C-reactive protein

In multivariable-adjusted models, we found that minoritized participants had 1.42 (95% CI: 1.20, 1.70) times higher odds of elevated CRP compared to non-Hispanic White participants. When each racialized group was analyzed separately, we found that non-Hispanic Black participants had 1.66 (95% CI: 1.33, 2.07) times higher odds of elevated CRP than their non-Hispanic White counterparts. This association was of lesser magnitude for Hispanic participants, who had only 1.14 (95% CI: 0.88, 1.47) times higher odds of elevated CRP than non-Hispanic White participants (**Supplemental Table 5**).

### Four-way mediation-interaction decomposition to assess C-reactive protein as a mediator of the racialized disparities in incident dementia

In our fully adjusted regression-based mediation models, the decomposition analysis comparing minoritized versus non-minoritized groups showed that the mediating effect of CRP accounted for 2% (95% CI: 0%, 6%) of the disparity in incident dementia, while the interaction effect between minoritized group status and elevated CRP accounted for 12% (95% CI: 2%, 22%) of the disparity (**Table 3 & Supplemental Figure 2**).

**Table 3:** 4-way mediation analysis decomposition for racialized disparities in incident dementia using elevated levels of C-reactive protein (CRP >4.57μg/mL) as mediator. Models are stratified by minoritized status and racialized social groups in a sample of United States adults in the Health and Retirement Study

| Mediator (CRP) | Incident Dementia* |  |  |  |  |  |  |  |  |  |  |  |
| --- | --- | --- | --- | --- | --- | --- | --- | --- | --- | --- | --- | --- |
|  | Minoritized Racial Group<br>vs non-Hispanic White |  |  |  | non-Hispanic Black<br>vs non-Hispanic White |  |  |  | Hispanic<br>vs non-Hispanic White |  |  |  |
|  | Estimate | [95%CI] | [95%CI] | p-value | Estimate | [95%CI] | [95%CI] | p-value | Estimate | [95%CI] | [95%CI] | p-value |
| **Excess Risk** |  |  |  |  |  |  |  |  |  |  |  |  |
| RERI Controlled Direct Effect | 1.91 | 1.54 | 2.38 | 0.00 | 2.37 | 1.76 | 3.13 | 0.00 | 1.46 | 0.97 | 2.09 | 0.00 |
| RERI Interaction Reference | 0.21 | 0.04 | 0.41 | 0.02 | 0.15 | -0.09 | 0.42 | 0.23 | 0.35 | 0.02 | 0.72 | 0.02 |
| RERI Interaction Mediation | 0.04 | 0.00 | 0.12 | 0.03 | 0.05 | -0.03 | 0.19 | 0.23 | -0.02 | -0.08 | 0.12 | 0.76 |
| RERI Pure Indirect Effect | 0.01 | 0.00 | 0.02 | 0.08 | 0.01 | 0.00 | 0.04 | 0.06 | 0.00 | -0.01 | 0.02 | 0.76 |
| **% Attributable** |  |  |  |  |  |  |  |  |  |  |  |  |
| % Controlled Direct Effect | 0.88 | 0.77 | 0.98 | 0.00 | 0.92 | 0.78 | 1.04 | 0.00 | 0.81 | 0.62 | 0.99 | 0.00 |
| % Interaction Reference | 0.10 | 0.02 | 0.19 | 0.02 | 0.06 | -0.03 | 0.15 | 0.23 | 0.20 | 0.01 | 0.37 | 0.02 |
| % Interaction Mediation | 0.02 | 0.00 | 0.05 | 0.03 | 0.02 | -0.01 | 0.07 | 0.23 | -0.01 | -0.04 | 0.06 | 0.76 |
| % Pure Indirect Effect | 0.00 | 0.00 | 0.01 | 0.08 | 0.01 | 0.00 | 0.02 | 0.06 | 0.00 | -0.01 | 0.01 | 0.76 |
| Percent Mediated | 0.02 | 0.00 | 0.06 | 0.02 | 0.02 | -0.01 | 0.08 | 0.13 | -0.01 | -0.05 | 0.07 | 0.76 |
| Percent due to Interaction | 0.12 | 0.02 | 0.22 | 0.02 | 0.08 | -0.05 | 0.21 | 0.23 | 0.19 | 0.01 | 0.38 | 0.02 |
| Percent Eliminated | 0.12 | 0.02 | 0.23 | 0.02 | 0.08 | -0.04 | 0.22 | 0.21 | 0.19 | 0.01 | 0.38 | 0.02 |

\*Outcome 6-year incident dementia, model adjusting for age, sex, education categories, *APOE-ε4* allele status, wave, smoking status, alcohol consumption, body mass index, and chronic conditions. Minoritized racial group: (non-Hispanic Black and Hispanic participants)

When decomposing the non-Hispanic Black vs non-Hispanic White disparity, we found that the mediating effect of CRP accounted for 2% (95% CI: -1%, 8%) of the disparity, and the portion attributable to the interaction accounted for 8% (95% CI: -5%, 21%) (**Table 3 & Supplemental Figure 2**). The Hispanic vs non-Hispanic White decomposition showed that the mediating effect of CRP was virtually zero. However, the portion attributable to the interaction accounted for 19% (95% CI: 1%, 38%) of the disparity (**Table 3 & Supplemental Figure 3**).

### Sensitivity Analysis

In this analysis, *APOE-ε4* may be a mediator-outcome confounder affected by our exposure of interest through racialized status being paired to geographic ancestry (**variable L, Figure 1**). Because of this potential violation of causal mediation analysis, we conducted a randomized analogue mediation model to test the robustness of our regression-based mediation estimates. We found that when comparing the minoritized group to the non-Hispanic White group, the mediating effect of CRP on incident dementia accounted for 3% (95% CI: 0%, 6%) of the disparity, and the proportion due to interaction accounted for 14% (95% CI: 2%, 25%) (**Supplemental Table 6 & Supplemental Figure 3**). When decomposing the non-Hispanic Black vs non-Hispanic White disparity, we found that the mediating effect of CRP accounted for 2% (95% CI: -1%, 8%) of the disparity, and the portion attributable to the interaction accounted for 8% (95% CI: -5%, 24%) (**Supplemental Table 6 & Supplemental Figure 3**). The Hispanic vs non-Hispanic White decomposition showed that the mediating effect of CRP accounted for 1% (95% CI: -5%, 7%), and the portion attributable to the interaction accounted for 22% (95% CI: 2%, 43%) of the disparity (**Supplemental Table 6 & Supplemental Figure 3**). These results were similar to those obtained from our regression-based estimates, suggesting that our findings were robust to a potential violation of mediation analysis. Moreover, our mediational E-value suggested that an unmeasured confounder associated with both high CRP and incident dementia with approximate rate ratios of 1.14-fold could completely explain away the observed indirect effect of the minoritized group vs. non-Hispanic White disparity, but a weaker confounder could not (**Supplemental Table 7A**). Further, an unmeasured confounder associated with both high CRP and incident dementia with approximate rate ratios of 1.05-fold could shift the mediated proportion confidence interval to the null, but a weaker confounder could not. Mediational E-values for the randomized analogue models were of slightly similar magnitude (**Supplement al Table 7B**).

## Discussion

Disparities in dementia among racialized groups are the result of multiple expressions of racism, and unveiling the biological mechanisms implicated in the production of these disparities is crucial for understanding how racism is embodied.^59^ In a nationally representative sample of older adults in the United States, we observed a 27% greater risk of incident dementia among those with high versus low CRP, and this association was stronger among Hispanic and non-Hispanic White participants than among non-Hispanic Black participants. We found that 12% of the observed disparity in incident dementia was accounted for by the interaction between minoritized group membership and elevated CRP, and 2% of the disparity was mediated by high CRP. A stronger interaction effect was apparent in the Hispanic versus non-Hispanic White decomposition, where we found that 19% of the disparity was attributable to the interaction effect between Hispanic group membership and high CRP. When decomposing the non-Hispanic Black versus non-Hispanic White disparity, we observed that 8% was attributable to the interaction effect between non-Hispanic Black membership and high CRP. These results indicate that systemic inflammation is associated with dementia risk, and the effect of high CRP on dementia is moderated by minoritized group status. When individuals are racialized as non-Hispanic Black and/or Hispanic, the effect of CRP on incident dementia risk is greater than expected had these individuals been racialized (and treated) as non-Hispanic White.^20^

Our findings fit with previous epidemiological studies describing differences in CRP levels across racialized groups.^29,34,37,60^ We found that non-Hispanic Black participants had higher circulating CRP than non-Hispanic White participants after adjusting for a wide range of covariates. These findings are consistent with those from another recent HRS analysis.^24^ Additionally, our results extend prior research linking systemic inflammation and dementia risk in large population-based studies. For instance, in a nested case-control study of Japanese American men (N=1,050), CRP levels of >1.0mg/L (vs. <0.34mg/L) were associated with 2.8 times greater odds of all dementia subtypes after adjusting for sociodemographic conditions, behavioral factors, and *APOE-ε4* carrier status.^34^ In a separate sample of community-dwelling older adults with a large number of non-Hispanic Black (N=1,255) and non-Hispanic White (N=1,776) participants, individuals in the highest tertile of CRP (2.5-85.2mg/L) had 1.41 greater odds of cognitive decline than participants in the lowest tertile (0.2-1.2mg/L), although no interaction effect between racialized group and inflammation was observed.^60^ Similarly, in a racially diverse sample of the Reasons for Geographic And Racial Differences in Stroke (REGARDS) cohort, average CRP levels were higher among Black participants (2.8mg/L; N=7,974) in comparison to their White counterparts (1.8mg/L; N=13,808).^29^ Using race-specific CRP cutoffs at the 90^th^ percentile, participants with baseline CRP at or above the 90^th^ percentile experienced a faster decline in memory and verbal fluency trajectories than those with CRP levels below the 90^th^ percentile.^29^ Again, in this study, researchers concluded that no interaction between racialized group and inflammation on cognition was present. Altogether, these prior studies suggest that elevated systemic inflammation is associated with adverse cognitive outcomes in older adults, and this effect was not modified by racialized groups. We expanded on these previous studies by incorporating a measure of additive interaction in our mediation models^61^ to test if the effect of high CRP levels on incident dementia was modified by the racialization process. This approach aligns with current epidemiological frameworks suggesting that effect modification is scale dependent, and the additive scale is better suited to test for interaction effects,^62^ and with more recent developments in mediation analysis that unify mediation and additive interaction into a unique framework.^61^ For instance, this innovative methodology allowed us to examine if the racialization process (implied in racialized group categories) modified the association between systemic inflammation and incident dementia, while simultaneously exploring whether systemic inflammation was a mediating pathway of the observed disparities.

Our results support the hypothesis that systemic inflammation is a plausible biological pathway implicated in the production of disparities in incident dementia. We found evidence that 2% of the disparity between the minoritized and privileged groups was attributable to the mediated CRP pathway, and another 12% was attributable to the moderated pathway. The slight mediation effect was expected since disparities between these groups emerge from structural forces acting differentially on groups rather than physiological processes that might be different among groups. These structural forces operate tacitly under the controlled direct effect, which represents a large proportion (88%) of the observed minoritized disparity. Systemic inflammation, and likely other biological responses, represent plausible mechanisms through which racism operates. The moderated pathway reflects the extent to which minoritized group status affects the association between CRP and incident dementia, which is greater than expected for individuals minoritized and racialized as non-Hispanic Black and/or Hispanic had these individuals been racialized and treated as non-Hispanic White. In other words, had all groups been treated comparably as non-Hispanic White individuals, disparities in incident dementia would be reduced.

Though there is complex reality when examining CRP as a biological pathway. In stratified mediation models, we observed weaker mediated and moderated effects when comparing the non-Hispanic Black versus non-Hispanic White disparity than when comparing the minoritized disparity. We attribute these findings to the weak association between CRP and incident dementia among non-Hispanic Black participants, for which we have two possible explanations. First, the small number of non-Hispanic Black participants could hinder our statistical power to detect a significant association. Second, we hypothesize that the higher levels of CRP found in non-Hispanic Black participants are characteristic of a chronic stress response that results from persistent experiences with structural racism.^16,63,64^ Therefore, chronic systemic inflammation may predispose Black participants for other competing events such as diabetes, cardiovascular disease, stroke, and premature death;^36,65,66^ which in turn may affect Black participants’ likelihood of retention during the study period. Although our models controlled for confounding bias by these potential competing events, we did not account for selection bias issues in our analysis, and future research should inform how differential loss to follow-up affects the relationship between systemic inflammation and dementia in Black participants. Additionally, we attribute the lack of mediation effect in the Hispanic vs non-Hispanic White disparity to the fact that differences in high CRP between these two groups were accounted for by individual-level confounders. However, we were able to detect important moderating effects for this disparity. Although we found consistent evidence that CRP was an important mediator of racialized disparities in dementia, our research was limited to a baseline measurement and a unique biomarker. Future research should incorporate multiple biomarkers of systemic inflammation and multiple time points to understand how trajectories of inflammation affect the cognitive function of older adults.

Our analysis has several strengths, including quantifying the association between CRP and incident dementia in a large (n=5,143) large and diverse sample of older adults in the United States. We had rich data on well-known confounding variables, including a major genetic risk factor for dementia. The prospective nature of our study design with 6 years of follow-up makes our results less susceptible to reverse causation. We also performed sensitivity analysis for our decomposition models and obtained consistent estimates.

Notably, treating *APOE-ε4* allele carrier status as a potential mediator-outcome confounder affected by the exposure did not alter our conclusions. We also tested the degree to which an unmeasured confounder could nullify our indirect effects by calculating mediational E-values for our decomposition models. Nonetheless, our major strength is the novel application of a recently developed methodological approach that unifies mediation and racialized category interaction effects into one scientific query.

The results of this study may serve as empirical evidence for existing theoretical frameworks that seek to explain how racism is embodied in the physiology of the individuals who live it, and how this embodiment affects their susceptibility to health and disease. The contextualization of race in causal methodology is part of an ongoing epidemiological debate. Our interpretation of disparities among racialized groups is up-to-date with recent developments on structural racism and causal methodology.^20,44^ Finally, this work has important implications for public health. We demonstrated that, in comparison to non-Hispanic White adults, minoritized groups in the United States have elevated levels of systemic inflammation even after controlling for individual-level factors. Therefore, public heath efforts should devote attention to understanding how structural racism and the process of racialization are associated with systemic inflammation in these populations, to ameliorate the racial gap in adverse cognitive outcomes.

## Data Availability

Survey data are publicly available, and genetic data are available through dbGaP and the National Institute on Aging Genetics of Alzheimer's Disease Data Storage Site

https://hrs.isr.umich.edu/data-products

https://dbgap.ncbi.nlm.nih.gov

https://dss.niagads.org/

## Acronyms

(*APOE*): apolipoprotein E
(CRP): C-reactive protein

## Acknowledgements

We thank the participants and staff of the Health and Retirement Study.

