## Supplemental Material for "The Mediating Role of Systemic Inflammation and Moderating Role of Race/Ethnicity in Racialized Disparities in Incident Dementia: A Decomposition Analysis"

**Supplemental Table 1:** Distribution of baseline characteristics between included and excluded participants in the United States Retirement Study

| Characteristic | **Overall** N = 8,781[1] | **Included** N = 5,143[1] | **Excluded** N = 3,638[1] | p-value[2] |
|---|---|---|---|---|
| **Cognitive Status** | | | | |
| Incident dementia | 795 (15%) | 795 (15%) | NA | |
| Cognitively normal | 4,348 (85%) | 4,348 (85%) | NA | |
| **Baseline CRP** (μg/mL) | 4.55 (8.14) | 4.18 (7.70) | 5.09 (8.69) | **<0.001** |
| **Age** (years) | 68.12 (10.38) | 66.36 (9.90) | 70.60 (10.52) | **<0.001** |
| **Race** | | | | **<0.001** |
| Non-Hispanic Black | 1,057 (12%) | 488 (9.5%) | 569 (16%) | |
| Hispanic | 794 (9.0%) | 376 (7.3%) | 418 (11%) | |
| Non-Hispanic Other | 189 (2.2%) | 0 (0%) | 189 (5.2%) | |
| Non-Hispanic White | 6,741 (77%) | 4,279 (83%) | 2,462 (68%) | |
| **Gender** | | | | **<0.001** |
| Female | 5,290 (60%) | 3,189 (62%) | 2,101 (58%) | |
| Male | 3,491 (40%) | 1,954 (38%) | 1,537 (42%) | |
| **Education Category** | | | | **<0.001** |
| > College | 812 (9.2%) | 602 (12%) | 210 (5.8%) | |
| College/Some | 1,575 (18%) | 1,109 (22%) | 466 (13%) | |
| HS or < | 6,394 (73%) | 3,432 (67%) | 2,962 (81%) | |
| **Alcohol** (# drinks/day when drinks) | 0.70 (1.41) | 0.74 (1.27) | 0.64 (1.59) | **0.002** |
| **Smoking** | | | | **0.005** |
| Current/Former | 4,950 (56%) | 2,835 (55%) | 2,115 (58%) | |
| Never | 3,831 (44%) | 2,308 (45%) | 1,523 (42%) | |
| **Body Mass Index** (kg/m$^2$) | 28.33 (5.88) | 28.46 (5.75) | 28.16 (6.05) | **0.018** |
| **Chronic Conditions** | | | | **<0.001** |
| At least 1 | 7,465 (85%) | 4,223 (82%) | 3,242 (89%) | |
| No | 1,316 (15%) | 920 (18%) | 396 (11%) | |
| ***APOE-ε4*** | | | | 0.190 |
| At least 1 | 2,373 (27%) | 1,363 (27%) | 1,010 (28%) | |
| No copy | 6,408 (73%) | 3,780 (73%) | 2,628 (72%) | |
| **Wave** | | | | **<0.001** |
| 2006 | 4,178 (48%) | 2,597 (50%) | 1,581 (43%) | |
| 2008 | 4,603 (52%) | 2,546 (50%) | 2,057 (57%) | |

[1]n (%); Mean (SD)

[2]Pearson's Chi-squared test; One-way ANOVA

CRP: C-reactive protein; *APOE-ε4*: apolipoprotein E *ε4* allele carrier status

**Supplemental Table 2:** Distribution of baseline sample characteristics by racialized social groups in the Health and Retirement Study, 2006 and 2008

| Characteristic | Overall Sample N = 5,143[1] | Non-Hispanic Black N = 488[1] | Hispanic N = 376[1] | Non-Hispanic White N = 4,279[1] | p-value[2] |
|---|---|---|---|---|---|
| **Dementia** | | | | | **<0.001** |
| Incident dementia | 795 (15%) | 171 (35%) | 103 (27%) | 521 (12%) | |
| Cognitively normal | 4,348 (85%) | 317 (65%) | 273 (73%) | 3,758 (88%) | |
| **Baseline CRP** (μg/mL) | 4.18 (7.70) | 6.19 (11.15) | 4.32 (6.29) | 3.94 (7.28) | **<0.001** |
| **Age** (years) | 66.36 (9.90) | 65.53 (9.68) | 63.63 (10.33) | 66.70 (9.85) | **<0.001** |
| **Sex** | | | | | **0.002** |
| Female | 3,189 (62%) | 338 (69%) | 232 (62%) | 2,619 (61%) | |
| Male | 1,954 (38%) | 150 (31%) | 144 (38%) | 1,660 (39%) | |
| **Educational Category** | | | | | **<0.001** |
| > College | 602 (12%) | 30 (6.1%) | 11 (2.9%) | 561 (13%) | |
| College/Some | 1,109 (22%) | 75 (15%) | 45 (12%) | 989 (23%) | |
| HS or < | 3,432 (67%) | 383 (78%) | 320 (85%) | 2,729 (64%) | |
| **Alcohol** (# drinks/day when drinks) | 0.74 (1.27) | 0.48 (1.14) | 0.65 (1.41) | 0.77 (1.27) | **<0.001** |
| **Smoking** | | | | | 0.496 |
| Current/Former | 2,835 (55%) | 279 (57%) | 200 (53%) | 2,356 (55%) | |
| Never | 2,308 (45%) | 209 (43%) | 176 (47%) | 1,923 (45%) | |
| **Body Mass Index** (kg/m$^2$) | 28.46 (5.75) | 30.69 (6.50) | 29.41 (5.50) | 28.12 (5.61) | **<0.001** |
| **Chronic Conditions** | | | | | **<0.001** |
| At least 1 | 4,223 (82%) | 432 (89%) | 291 (77%) | 3,500 (82%) | |
| No | 920 (18%) | 56 (11%) | 85 (23%) | 779 (18%) | |
| ***APOE-ε4*** | | | | | **<0.001** |
| At least 1 copy | 1,363 (27%) | 185 (38%) | 83 (22%) | 1,095 (26%) | |
| No copy | 3,780 (73%) | 303 (62%) | 293 (78%) | 3,184 (74%) | |
| **Time** (years) | | | | | **<0.001** |
| 2 | 302 (5.9%) | 80 (16%) | 39 (10%) | 183 (4.3%) | |
| 4 | 269 (5.2%) | 52 (11%) | 37 (9.8%) | 180 (4.2%) | |
| 6 | 4,572 (89%) | 356 (73%) | 300 (80%) | 3,916 (92%) | |

| **Characteristic** | **Overall Sample** N = 5,143[1] | **Non-Hispanic Black** N = 488[1] | **Hispanic** N = 376[1] | **Non-Hispanic White** N = 4,279[1] | p-value[2] |
|---|---|---|---|---|---|
| **Wave** | | | | | **0.017** |
| 2006 | 2,597 (50%) | 224 (46%) | 174 (46%) | 2,199 (51%) | |
| 2008 | 2,546 (50%) | 264 (54%) | 202 (54%) | 2,080 (49%) | |

[1]n (%); Mean (SD)

[2]Pearson's Chi-squared test; One-way ANOVA

CRP: C-reactive protein; *APOE-ε4*: apolipoprotein E *ε4* allele carrier status

**Supplemental Table 3:** Baseline levels of C-reactive protein (CRP) by racialized social groups and sex in the United States Health and Retirement Study, waves 2006 & 2008

| Race/ethnicity | Sex | Observations | Mean | SD | Min | Max | 25th | 50th | 75th | IQR |
|---|---|---|---|---|---|---|---|---|---|---|
| **Non-Hispanic Black** | **Female** | 338 | 6.28 | 9.04 | 0.04 | 81.1 | 1.42 | 3.60 | 7.77 | 6.34 |
| **Non-Hispanic Black** | **Male** | 150 | 6.00 | 14.9 | 0.03 | 125.0 | 1.14 | 2.49 | 5.86 | 4.72 |
| **Hispanic** | **Female** | 232 | 4.61 | 5.59 | 0.08 | 37.9 | 1.40 | 2.50 | 5.79 | 4.39 |
| **Hispanic** | **Male** | 144 | 3.85 | 7.28 | 0.04 | 66.7 | 0.99 | 1.89 | 4.04 | 3.06 |
| **Non-Hispanic White** | **Female** | 2619 | 4.27 | 7.57 | 0.04 | 173.0 | 1.01 | 2.13 | 4.91 | 3.90 |
| **Non-Hispanic White** | **Male** | 1660 | 3.40 | 6.77 | 0.02 | 121.0 | 0.79 | 1.55 | 3.30 | 2.51 |

**Supplemental Table 4:** Distribution of baseline sample characteristics by percentiles of C-reactive protein, United States Health and Retirement Study, 2006 and 2008

| | | **C-reactive protein (μg/mL) percentiles** | | | | |
|---|---|---|---|---|---|---|
| **Characteristic** | **Overall** N = 5,143[1] | **$25^{th}$ (<0.96)** N = 1,282[1] | **$25^{th}$ - $50^{th}$ (≥0.96 & <1.97)** N = 1,267[1] | **$50^{th}$ - $75^{th}$ (≥1.97 & <4.57)** N = 1,306[1] | **$75^{th}$ (≥4.57)** N = 1,288[1] | p-value[2] |
| **Dementia Status** | | | | | | 0.115 |
| Incident dementia | 795 (15%) | 195 (15%) | 177 (14%) | 199 (15%) | 224 (17%) | |
| Cognitively normal | 4,348 (85%) | 1,087 (85%) | 1,090 (86%) | 1,107 (85%) | 1,064 (83%) | |
| **Age** (years) | 66.36 (9.90) | 66.35 (10.17) | 67.05 (9.77) | 66.60 (9.73) | 65.45 (9.88) | **<0.001** |
| **Race** | | | | | | **<0.001** |
| Non-Hispanic Black | 488 (9.5%) | 88 (6.9%) | 75 (5.9%) | 133 (10%) | 192 (15%) | |
| Hispanic | 376 (7.3%) | 71 (5.5%) | 91 (7.2%) | 107 (8.2%) | 107 (8.3%) | |
| Non-Hispanic White | 4,279 (83%) | 1,123 (88%) | 1,101 (87%) | 1,066 (82%) | 989 (77%) | |
| **Sex** | | | | | | **<0.001** |
| Female | 3,189 (62%) | 718 (56%) | 719 (57%) | 836 (64%) | 916 (71%) | |
| Male | 1,954 (38%) | 564 (44%) | 548 (43%) | 470 (36%) | 372 (29%) | |
| **Educational Category** | | | | | | **<0.001** |
| > College | 602 (12%) | 221 (17%) | 148 (12%) | 124 (9.5%) | 109 (8.5%) | |
| College/Some | 1,109 (22%) | 297 (23%) | 308 (24%) | 261 (20%) | 243 (19%) | |
| HS or < | 3,432 (67%) | 764 (60%) | 811 (64%) | 921 (71%) | 936 (73%) | |
| **Alcohol** (# drinks/day when drinks) | 0.74 (1.27) | 0.83 (1.27) | 0.81 (1.33) | 0.71 (1.26) | 0.60 (1.21) | **<0.001** |
| **Smoking Status** | | | | | | **<0.001** |
| Current/Former | 2,835 (55%) | 631 (49%) | 708 (56%) | 743 (57%) | 753 (58%) | |
| Never | 2,308 (45%) | 651 (51%) | 559 (44%) | 563 (43%) | 535 (42%) | |
| **Body Mass Index** ($kg/m^2$) | 28.46 (5.75) | 25.89 (4.30) | 27.48 (4.66) | 29.01 (5.34) | 31.40 (6.84) | **<0.001** |
| **Chronic Conditions** | | | | | | **<0.001** |
| At least 1 | 4,223 (82%) | 972 (76%) | 1,008 (80%) | 1,114 (85%) | 1,129 (88%) | |

| **Characteristic** | **Overall**<br>N = 5,143[1] | **C-reactive protein (μg/mL) percentiles**<br>**25th**<br>**(<0.96)**<br>N = 1,282[1] | <br>**25th - 50th**<br>**(≥0.96 & <1.97)**<br>N = 1,267[1] | <br>**50th - 75th**<br>**(≥1.97 & <4.57)**<br>N = 1,306[1] | <br>**75th**<br>**(≥4.57)**<br>N = 1,288[1] | p-value[2] |
|---|---|---|---|---|---|---|
| No | 920 (18%) | 310 (24%) | 259 (20%) | 192 (15%) | 159 (12%) | |
| ***APOE-ε4*** | | | | | | **<0.001** |
| At least 1 copy | 1,363 (27%) | 447 (35%) | 347 (27%) | 301 (23%) | 268 (21%) | |
| No copy | 3,780 (73%) | 835 (65%) | 920 (73%) | 1,005 (77%) | 1,020 (79%) | |
| **Time** (years) | | | | | | **0.009** |
| 2 | 302 (5.9%) | 58 (4.5%) | 80 (6.3%) | 68 (5.2%) | 96 (7.5%) | |
| 4 | 269 (5.2%) | 71 (5.5%) | 51 (4.0%) | 80 (6.1%) | 67 (5.2%) | |
| 6 | 4,572 (89%) | 1,153 (90%) | 1,136 (90%) | 1,158 (89%) | 1,125 (87%) | |
| **Wave** | | | | | | 0.470 |
| 2006 | 2,597 (50%) | 659 (51%) | 640 (51%) | 636 (49%) | 662 (51%) | |
| 2008 | 2,546 (50%) | 623 (49%) | 627 (49%) | 670 (51%) | 626 (49%) | |

[1]n (%); Mean (SD)

[2]Pearson's Chi-squared test; One-way ANOVA

*APOE-ε4*: apolipoprotein E *ε4* allele carrier status

**Supplemental Table 5:** Odds ratio of elevated levels of C-reactive protein ($\geq$4.57μg/mL or $\geq$ $75^{th}$ percentile) stratified by minoritized status and racialized social groups in the US Health and Retirement Study, waves 2006 & 2008

| | **Non-Hispanic Black vs non-Hispanic White[1]** N = 4,767 | | **Hispanic vs non-Hispanic White[1]** N = 4,655 | | **Minoritized Racial Group vs non-Hispanic White[2]** N = 5,143 | |
|---|---|---|---|---|---|---|
| **Models** | | | | | | |
| **Unadjusted** | $2.16^{***}$ | [1.78,2.62] | $1.32^{*}$ | [1.05,1.67] | $1.76^{***}$ | [1.50,2.06] |
| **Demographic[+]** | $2.11^{***}$ | [1.72,2.58] | 1.17 | [0.92,1.49] | $1.66^{***}$ | [1.41,1.95] |
| **Risk Factors[$]** | $1.67^{***}$ | [1.34,2.08] | 1.13 | [0.87,1.46] | $1.42^{***}$ | [1.20,1.70] |
| **Chronic Conditions[§]** | $1.66^{***}$ | [1.33,2.07] | 1.14 | [0.88,1.47] | $1.42^{***}$ | [1.20,1.70] |

Demographic model: adjusted for age, sex, education categories, *APOE-ε4* allele status, and wave

[$] Risk factors model: adjusted for age, sex, education categories, *APOE-ε4* allele status, wave, smoking status, alcohol consumption, body mass index

[§] Chronic conditions model: adjusted for age, sex, education categories, *APOE-ε4* allele status, wave, smoking status, alcohol consumption, body mass index, and chronic conditions

[1] Odds ratios, CI: confidence interval in brackets

[2] Odds ratios, CI: confidence intervals in brackets. Minoritized racial group: (non-Hispanic Black and Hispanic participants)

$^{*}$ $p < 0.05$, $^{**}$ $p < 0.01$, $^{***}$ $p < 0.001$

**Supplemental Table 6:** Randomized analogue mediation models for racial disparities in incident dementia using elevated levels of C-reactive protein (CRP $\geq$4.57μg/mL) as mediator. Models are stratified by minoritized status and racialized social groups in a sample of United States adults in the Health and Retirement Study

| | Randomized Analogue Model+ (Outcome: Incident Dementia) | | | | | | | | | | | |
|---|---|---|---|---|---|---|---|---|---|---|---|---|
| | Minoritized Racial Group vs non-Hispanic White N = 5,143 | | | | non-Hispanic Black vs non-Hispanic White N = 4,767 | | | | Hispanic vs non-Hispanic White N = 4,655 | | | |
| **Mediator (CRP)** | **Estimate** | **[95%CI]** | **[95%CI]** | **p-value** | **Estimate** | **[95%CI]** | **[95%CI]** | **p-value** | **Estimate** | **[95%CI]** | **[95%CI]** | **p-value** |
| **Excess Risk** | | | | | | | | | | | | |
| RERI Controlled Direct Effect | 1.94 | 1.58 | 2.44 | <0.00 | 2.57 | 1.90 | 3.32 | <0.00 | 1.40 | 0.94 | 2.00 | <0.00 |
| RERI Interaction Reference | 0.26 | 0.04 | 0.52 | 0.02 | 0.18 | -0.11 | 0.53 | 0.22 | 0.39 | 0.03 | 0.87 | 0.03 |
| RERI Interaction Mediation | 0.06 | -0.01 | 0.13 | 0.08 | 0.05 | -0.03 | 0.21 | 0.23 | 0.01 | -0.09 | 0.14 | 0.68 |
| RERI Pure Indirect Effect | 0.01 | 0.00 | 0.02 | 0.12 | 0.01 | 0.00 | 0.04 | 0.07 | 0.00 | -0.01 | 0.02 | 0.71 |
| **% Attributable** | | | | | | | | | | | | |
| % Controlled Direct Effect | 0.85 | 0.75 | 0.97 | <0.00 | 0.91 | 0.76 | 1.05 | <0.00 | 0.78 | 0.57 | 0.98 | <0.00 |
| % Interaction Reference | 0.11 | 0.02 | 0.22 | 0.02 | 0.06 | -0.04 | 0.18 | 0.22 | 0.22 | 0.02 | 0.42 | 0.03 |
| % Interaction Mediation | 0.03 | 0.00 | 0.05 | 0.08 | 0.02 | -0.01 | 0.07 | 0.23 | 0.00 | -0.05 | 0.07 | 0.68 |
| % Pure Indirect Effect | 0.01 | 0.00 | 0.01 | 0.12 | 0.00 | 0.00 | 0.01 | 0.07 | 0.00 | -0.01 | 0.01 | 0.71 |
| Percent Mediated | 0.03 | 0.00 | 0.06 | 0.07 | 0.02 | -0.01 | 0.08 | 0.13 | 0.01 | -0.05 | 0.07 | 0.68 |
| Percent due to Interaction | 0.14 | 0.02 | 0.25 | 0.02 | 0.08 | -0.05 | 0.24 | 0.22 | 0.22 | 0.02 | 0.43 | 0.03 |
| Percent Eliminated | 0.15 | 0.03 | 0.25 | 0.01 | 0.09 | -0.05 | 0.24 | 0.21 | 0.22 | 0.02 | 0.43 | 0.03 |

+Model: outcome 6-year incident dementia, model adjusting for age, sex, education categories, wave, smoking status, alcohol consumption, body mass index, alcohol consumption, and chronic conditions, using *APOE-ε4* as variable affected by the race/ethnicity through ancestry.
Minoritized racial group: (non-Hispanic Black and Hispanic participants)

**Supplemental Table 7A:** Mediational E-values for regression-based models; estimates are presented in the rate ratio scale. Models are stratified by racialized social groups in a sample of the United States adults in the Health and Retirement Study.

| | Minoritized vs non-Hispanic White* | | | | | |
|---|---|---|---|---|---|---|
| **Rate Ratio Scale** | **Estimate** | **LB 95%CI** | **UB 95%CI** | **E-value** | **E-value LB** | **E-value UB** |
| Controlled direct effect (Rcde) | 2.99 | 2.58 | 3.50 | 5.43 | 4.61 | NA |
| Pure natural direct effect (Rpnde) | 3.13 | 2.75 | 3.58 | 5.71 | 4.95 | NA |
| Total natural direct effect (Rtnde) | 3.15 | 2.78 | 3.62 | 5.76 | 5.00 | NA |
| Pure natural indirect effect (Rpnie) | 1.01 | 1.00 | 1.02 | 1.09 | 1.00 | NA |
| Total natural indirect effect (Rtnie) | 1.02 | 1.00 | 1.04 | 1.14 | 1.05 | NA |
| Total effect (Rte) | 3.17 | 2.79 | 3.66 | 5.80 | 5.03 | NA |
| | **non-Hispanic Black vs non-Hispanic White*** | | | | | |
| | **Estimate** | **LB 95%CI** | **UB 95%CI** | **E-value** | **E-value LB** | **E-value UB** |
| Controlled direct effect (Rcde) | 3.48 | 2.84 | 4.26 | 6.42 | 5.12 | NA |
| Pure natural direct effect (Rpnde) | 3.53 | 2.96 | 4.23 | 6.51 | 5.36 | NA |
| Total natural direct effect (Rtnde) | 3.54 | 2.98 | 4.25 | 6.54 | 5.41 | NA |
| Pure natural indirect effect (Rpnie) | 1.01 | 1.00 | 1.04 | 1.13 | 1.00 | NA |
| Total natural indirect effect (Rtnie) | 1.02 | 0.99 | 1.06 | 1.15 | 1.00 | NA |
| Total effect (Rte) | 3.59 | 3.04 | 4.30 | 6.64 | 5.53 | NA |
| | **Hispanic vs non-Hispanic White*** | | | | | |
| | **Estimate** | **LB 95%CI** | **UB 95%CI** | **E-value** | **E-value LB** | **E-value UB** |
| Controlled direct effect (Rcde) | 2.53 | 2.02 | 3.19 | 4.50 | 3.45 | NA |
| Pure natural direct effect (Rpnde) | 2.82 | 2.33 | 3.45 | 5.08 | 4.09 | NA |
| Total natural direct effect (Rtnde) | 2.81 | 2.35 | 3.46 | 5.06 | 4.13 | NA |
| Pure natural indirect effect (Rpnie) | 1.00 | 0.99 | 1.02 | 1.04 | NA | 1.00 |
| Total natural indirect effect (Rtnie) | 0.99 | 0.97 | 1.04 | 1.08 | NA | 1.00 |
| Total effect (Rte) | 2.80 | 2.35 | 3.47 | 5.05 | 4.14 | NA |

* These E-values are on the rate ratio scale and correspond to the decomposition presented on Table 3
LB: Lower bound; UB: Upper bound
95%CI: 95% Confidence interval
NA: not applicable

**Supplemental Table 7B:** Mediational E-values for randomized analogue models; estimates are presented in the rate ratio scale. Models are stratified by racialized social groups in a sample of the United States adults in the Health and Retirement Study.

| | | | Minoritized vs non-Hispanic White* | | | |
|---|---|---|---|---|---|---|
| **Rate Ratio Scale** | **Estimate** | **LB 95%CI** | **UB 95%CI** | **E-value** | **E-value LB** | **E-value UB** |
| Controlled direct effect (Rcde) | 3.03 | 2.65 | 3.58 | 5.51 | 4.74 | NA |
| Pure natural direct effect (rRpnde)+ | 3.20 | 2.83 | 3.71 | 5.85 | 5.10 | NA |
| Total natural direct effect (rRtnde)+ | 3.23 | 2.86 | 3.77 | 5.92 | 5.16 | NA |
| Pure natural indirect effect (rRpnie)+ | 1.01 | 1.00 | 1.02 | 1.12 | 1.00 | NA |
| Total natural indirect effect (rRtnie)+ | 1.02 | 1.00 | 1.04 | 1.18 | 1.00 | NA |
| Total effect (Rte) | 3.27 | 2.88 | 3.80 | 6.00 | 5.21 | NA |
| | | | **non-Hispanic Black vs non-Hispanic White*** | | | |
| | **Estimate** | **LB 95%CI** | **UB 95%CI** | **E-value** | **E-value LB** | **E-value UB** |
| Controlled direct effect (Rcde) | 3.69 | 2.99 | 4.50 | 6.85 | 5.43 | NA |
| Pure natural direct effect (rRpnde)+ | 3.75 | 3.15 | 4.46 | 6.95 | 5.76 | NA |
| Total natural direct effect (rRtnde)+ | 3.76 | 3.17 | 4.51 | 6.98 | 5.78 | NA |
| Pure natural indirect effect (rRpnie)+ | 1.01 | 1.00 | 1.04 | 1.13 | 1.00 | NA |
| Total natural indirect effect (rRtnie)+ | 1.02 | 0.99 | 1.06 | 1.15 | 1.00 | NA |
| Total effect (Rte) | 3.81 | 3.22 | 4.56 | 7.09 | 5.90 | NA |
| | | | **Hispanic vs non-Hispanic White*** | | | |
| | **Estimate** | **LB 95%CI** | **UB 95%CI** | **E-value** | **E-value LB** | **E-value UB** |
| Controlled direct effect (Rcde) | 2.47 | 1.97 | 3.12 | 4.38 | 3.36 | NA |
| Pure natural direct effect (rRpnde)+ | 2.79 | 2.32 | 3.48 | 5.03 | 4.08 | NA |
| Total natural direct effect (rRtnde)+ | 2.80 | 2.33 | 3.50 | 5.04 | 4.09 | NA |
| Pure natural indirect effect (rRpnie)+ | 1.00 | 0.99 | 1.02 | 1.01 | 1.00 | NA |
| Total natural indirect effect (rRtnie)+ | 1.00 | 0.97 | 1.05 | 1.06 | 1.00 | NA |
| Total effect (Rte) | 2.80 | 2.35 | 3.52 | 5.04 | 4.12 | NA |

* These E-values are on the rate ratio scale and correspond to the decomposition presented on Supplemental Table 6
+ Randomized analogue estimate
LB: Lower bound; UB: Upper bound
95%CI: 95% Confidence interval
NA: not applicable

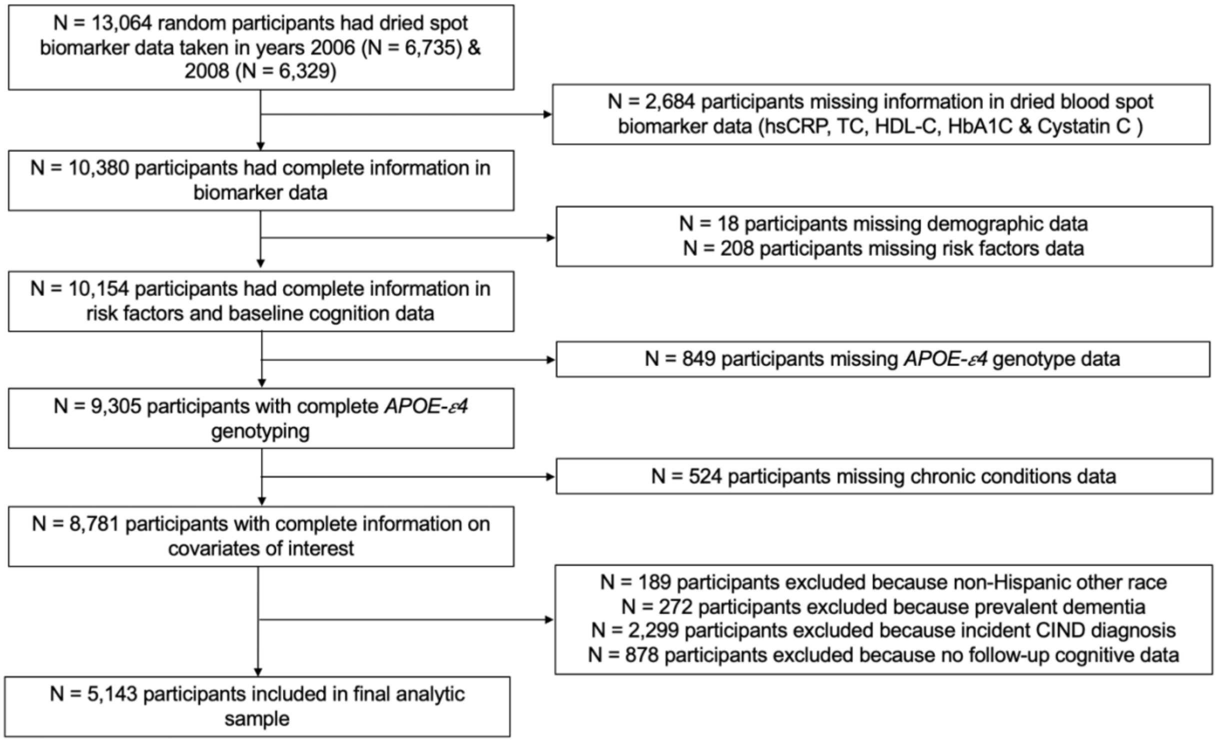


**Supplemental Figure 1:** Flow diagram of analytic sample study in the Health and Retirement Study. CRP: C-reactive protein, TC: total cholesterol; HDL-C: high density lipoprotein; HbA1C: glycosylated hemoglobin, *APOE-ε4*: apolipoprotein E *ε4* allele carrier status, CIND: cognitive impairment non-dementia.

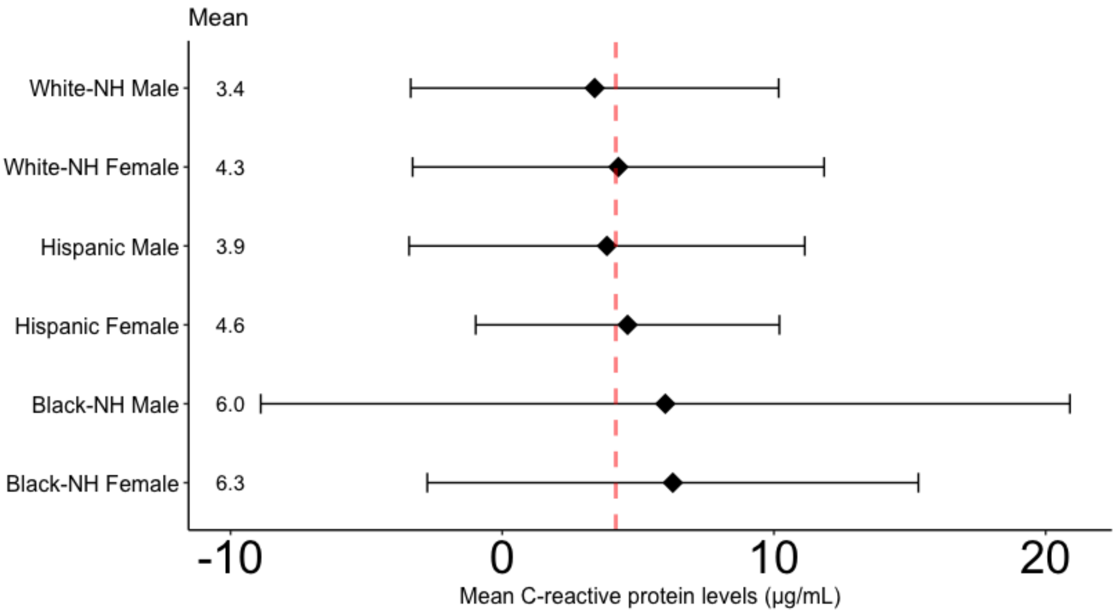


**Supplemental Figure 2:** Baseline C-reactive protein levels (μg/mL) by racialized social groups and sex in the United States Health and Retirement Study, waves 2006 & 2008. Dotted red line denotes the average levels of C-reactive protein in the overall sample (4.18μg/mL). White-NH: non-Hispanic White, Black-NH: non-Hispanic Black.

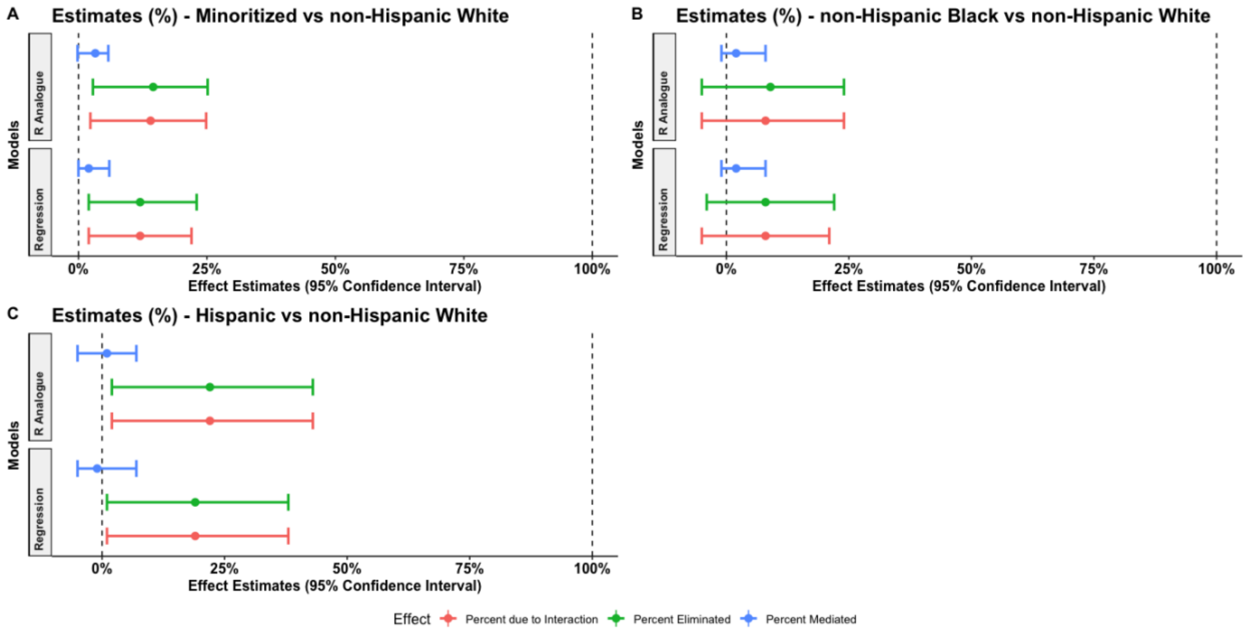


**Supplemental Figure 3:** Plot of mediation analysis estimates from regression-based and randomized analogue models denoting percent of the racial disparity in incident dementia that is due to the mediating effect of C-reactive protein, the percent due to the interaction between exposure and mediator, and the proportion eliminated. **A.** Mediation estimates from the racial disparity between the minoritized racial group (non-Hispanic Black and Hispanic) vs the non-Hispanic White group. **B.** Mediation estimates from the racial disparity between the non-Hispanic Black vs the non-Hispanic White group. **C.** Mediation estimates from the racial disparity between the Hispanic group vs the non-Hispanic White group.

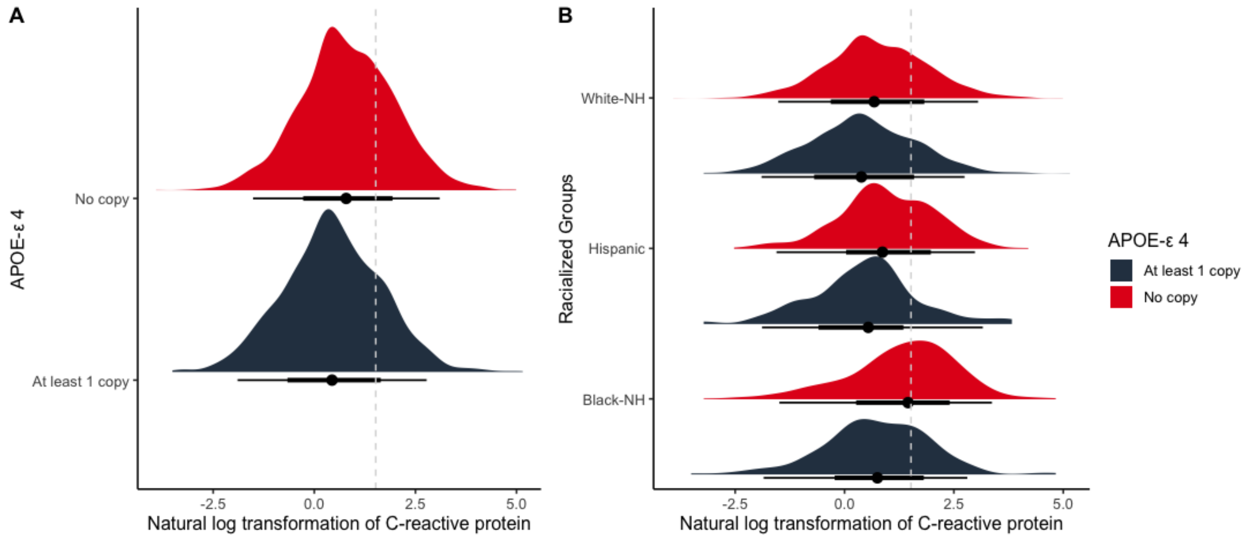


**Supplemental Figure 4:** Density plot of natural logarithmic transformation of C-reactive protein (CRP) in a selected sample of United States adults in the Health and Retirement Study.
**A.** Distribution of the natural logarithmic transformation of C-reactive protein (CRP) by *APOE-ε4* allele carrier status in our selected sample from the Health and Retirement Study (HRS). Dotted line denotes the cut off point for elevated levels of CRP at the 75th percentile (≥ 4.57μg/mL).
**B.** Distribution of CRP by racialized groups and *APOE-ε4* allele carrier status. White-NH: non-Hispanic White, Black-NH: non-Hispanic Black.
